# Real-life clinical sensitivity of SARS-CoV-2 RT-PCR test in symptomatic patients

**DOI:** 10.1101/2020.11.01.20223107

**Authors:** Elisa Kortela, Vesa Kirjavainen, Maarit J. Ahava, Suvi T. Jokiranta, Anna But, Anna Lindahl, Anu E. Jääskeläinen, Annemarjut J. Jääskeläinen, Asko Järvinen, Pia Jokela, Hannimari Kallio-Kokko, Raisa Loginov, Laura Mannonen, Eeva Ruotsalainen, Tarja Sironen, Olli Vapalahti, Maija Lappalainen, Hanna-Riikka Kreivi, Hanna Jarva, Satu Kurkela, Eliisa Kekäläinen

**Affiliations:** Division of Infectious diseases, Inflammation Center, Helsinki University Hospital and University of Helsinki, Helsinki, Finland; HUSLAB Clinical Microbiology, HUS Diagnostic Center, University of Helsinki and Helsinki University Hospital, Finland; Translational Immunology Research Program and Department of Bacteriology and Immunology, University of Helsinki, Helsinki, Finland; Biostatistics consulting, Department of Public Health, University of Helsinki and Helsinki University Hospital, Helsinki, Finland; Department of Respiratory Medicine, Heart and Lung Center, Helsinki University Hospital, University of Helsinki, Helsinki, Finland; Department of Virology, Faculty of Medicine, University of Helsinki, Helsinki, Finland; Department of Veterinary Biosciences, Faculty of Veterinary Medicine, University of Helsinki, Helsinki, Finland

## Abstract

**Importance:** Understanding the false negative rates of SARS-CoV-2 RT-PCR testing is pivotal for the management of the COVID-19 pandemic and it has practical implications for patient management in healthcare facilities.

**Objective:** To determine the real-life clinical sensitivity of SARS-CoV-2 RT-PCR testing.

**Design:** A retrospective study on case series from 4 March – 15 April 2020.

**Setting:** A population-based study conducted in primary and tertiary care in the Helsinki Capital Region, Finland.

**Participants:** Adults who were clinically suspected of SARS-CoV-2 infection and underwent SARS-CoV-2 RT-PCR testing, and who had sufficient data for grading of clinical suspicion of COVID-19 in their medical records were eligible. All 1,194 inpatients admitted to COVID-19 cohort wards during the study period were included. The outpatient cohort of 1,814 individuals was sampled from epidemiological line lists by systematic quasi-random sampling. Altogether 83 eligible outpatients (4.6%) and 3 inpatients (0.3%) were excluded due to insufficient data for grading of clinical suspicion.

**Exposures:** High clinical suspicion for COVID-19 was used as the reference standard for the RT-PCR test. Patients were considered to have high clinical suspicion of COVID-19 if the physician in charge recorded the suspicion on clinical grounds, or the patient fulfilled specifically defined clinical and exposure criteria.

**Main measures:** Sensitivity of SARS-CoV-2 RT-PCR by using manually curated clinical characteristics as the gold standard.

**Results:** The study population included 1,814 outpatients (mean [SD] age, 45.4 [17.2] years; 69.1% women) and 1,194 inpatients (mean [SD] age, 63.2 [18.3] years; 45.2% women). The sensitivity (95% CI) for laboratory confirmed cases, i.e. repeatedly tested patients were as follows: 85.7% (81.5–89.1%) inpatients; 95.5% (92.2–97.5%) outpatients, 89.9% (88.2–92.1%) all. When also patients that were graded as high suspicion but never tested positive were included in the denominator, the following sensitivity values (95% CI) were observed: 67.5% (62.9–71.9%) inpatients; 34.9% (31.4–38.5%) outpatients; 47.3% (44.4–50.3%) all.

**Conclusions and relevance:** The clinical sensitivity of SARS-CoV-2 RT-PCR testing was only moderate at best. The relatively high false negative rates of SARS-CoV-2 RT-PCR testing need to be accounted for in clinical decision making, epidemiological interpretations and when using RT-PCR as a reference for other tests.

**Key Points:** *Question:* What is the clinical sensitivity of SARS-CoV-2 RT-PCR test?

*Findings:* In this population-based retrospective study on medical records of 1,814 outpatients and 1,194 inpatients, the clinical sensitivity of SARS-CoV-2 RT-PCR was 47.3–89.9%.

*Meaning:* The false negative rates of SARS-CoV-2 RT-PCR testing need to be accounted for in clinical decision making, epidemiological interpretations and when using RT-PCR as a reference for other tests.

## Introduction

During the COVID-19 pandemic a central method for limiting the spread of SARS-CoV-2 has been the so-called “Test, Trace, Isolate” (TTI) approach promoted by the World Health Organization.^1-2^ A key feature of any laboratory test is its efficacy in detecting true positive cases. Evidence suggests a fair analytical sensitivity for the SARS-CoV-2 RT-PCR tests available on the market.^3-4^ However, recent reports indicate that clinically evident COVID-19 infections often go undetected by SARS-CoV-2 RT-PCR testing.^5-9^

A number of pivotal factors may decrease the overall sensitivity of testing and its usefulness in the TTI strategy. Preanalytical pitfalls such as suboptimal specimen collection may affect sample quality and hamper test sensitivity. Variation in viral shedding in different anatomical locations, and temporal variation in relation to disease onset can influence detection rates.^10-11^

High false negative rate complicates controlling the epidemic but it also has implications for healthcare settings.^12^ Removal of infection control precautions in hospitalized patients due to a false negative test causes an occupational hazard for healthcare workers and can lead to nosocomial spread of the disease. Real-life sensitivity estimates in the initial reports^13-14^ were limited by small sample sizes and variable testing methods and reference standards.

We decided to evaluate the clinical sensitivity of SARS-CoV-2 RT-PCR in a population-based setting in the beginning of the epidemic with low level of transmission. To avoid bias created by the high pretest probability of inpatients, our analysis also included outpatients. We used manually curated clinical characteristics from a cohort of 3,008 individuals as the gold standard for the RT-PCR test. We focused on the sensitivity of the first SARS-CoV-2 RT-PCR test since for outpatients repeated testing is not often feasible. Our analysis that uses only non-dependent samples will provide a more accurate estimate of clinical sensitivity since it is not confounded by bias from repeated sampling of the same individuals. Our data can be directly used to inform the practicing clinicians and epidemiologists how well the RT-PCR performs in a low prevalence setting.

## Materials and methods

### Study design and participants

We present data from a retrospective study conducted from electronic, comprehensive medical records. The study complies with the STARD reporting guidelines^15^, and it was approved by the local review board (HUS/157/2020-29).

During the study period 4 March – 15 April 2020, 22,821 individuals underwent SARS-CoV-2 RT-PCR testing at HUSLAB laboratory, Helsinki University Hospital, Finland, which serves the Helsinki Capital Area in Finland. On 31 March, the cumulative number of COVID-19 cases was 855, incidence in the previous two weeks was 44.3/100,000 according to the National Institute of Health and Welfare Finland, and there were 100 inpatients. On 15 April, the cumulative number of cases was 2,133, incidence in previous two weeks was 74.0/100,000, and there were 159 inpatients. eFigure 1 shows the number of daily specimens and proportion of positive specimens at HUSLAB during the study period.

We reasoned that the pretest probability, i.e. the probability for testing positive, would be different for inpatients on the COVID-19 cohort wards and outpatients and decided to study these two populations separately.

#### Outpatient cohort

During the study period, the tested patients were generally symptomatic but the criteria which prompted testing varied slightly over time (eTable 1). Initially, persons returning from recognized epidemic areas and exhibiting respiratory symptoms within 14 days of return were primarily tested. The criteria were soon expanded to include symptomatic persons with risk factors, and all symptomatic healthcare workers. Outpatients fulfilling testing criteria were recorded manually with some clinical details on a line list. These lists were the most systematically collected dataset for the tested outpatients so we chose to sample our outpatient cohort from these lists. We performed systematic (quasi-random) sampling by including every fifth individual from the line lists. Along with practical advantages, this approach decreased the probability of sampling dependent individuals, such as members of the same family. Besides the clinical details on the line lists, we checked electronic medical records for comorbidities and other demographic details. Other exclusion criteria for outpatients were age below 18 years and residence outside of the Helsinki and Uusimaa Hospital District. Altogether 1,814 eligible outpatients were included in the study (Figure 1A).

**Figure 1A.**
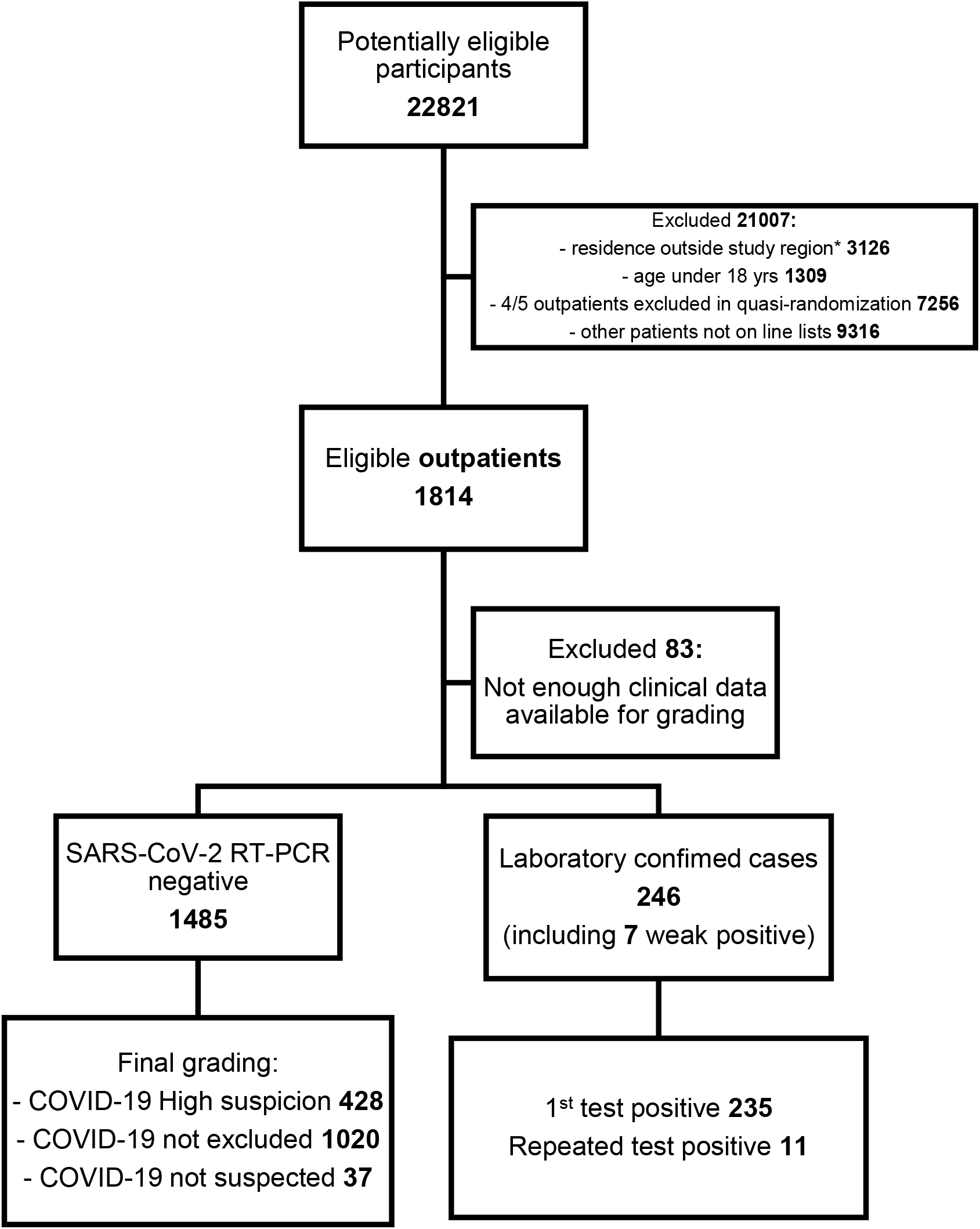
Selection of the outpatient cohort presented as a flowchart.

#### Inpatient cohort

Patients with fever, respiratory or gastrointestinal symptoms, and/or difficulty in breathing were suspected for COVID-19 and treated in designated cohort wards: 11 wards and 6 ICUs in eight hospitals (list of wards in eTable 2). All patients aged >18 years admitted to one of the cohort wards were eligible for the study and only patients without a SARS-CoV-2 RT-PCR performed at the HUSLAB laboratory were excluded. These inpatients formed a consecutive case series of 1,194 individuals (Figure 1B).

**Figure 1B.**
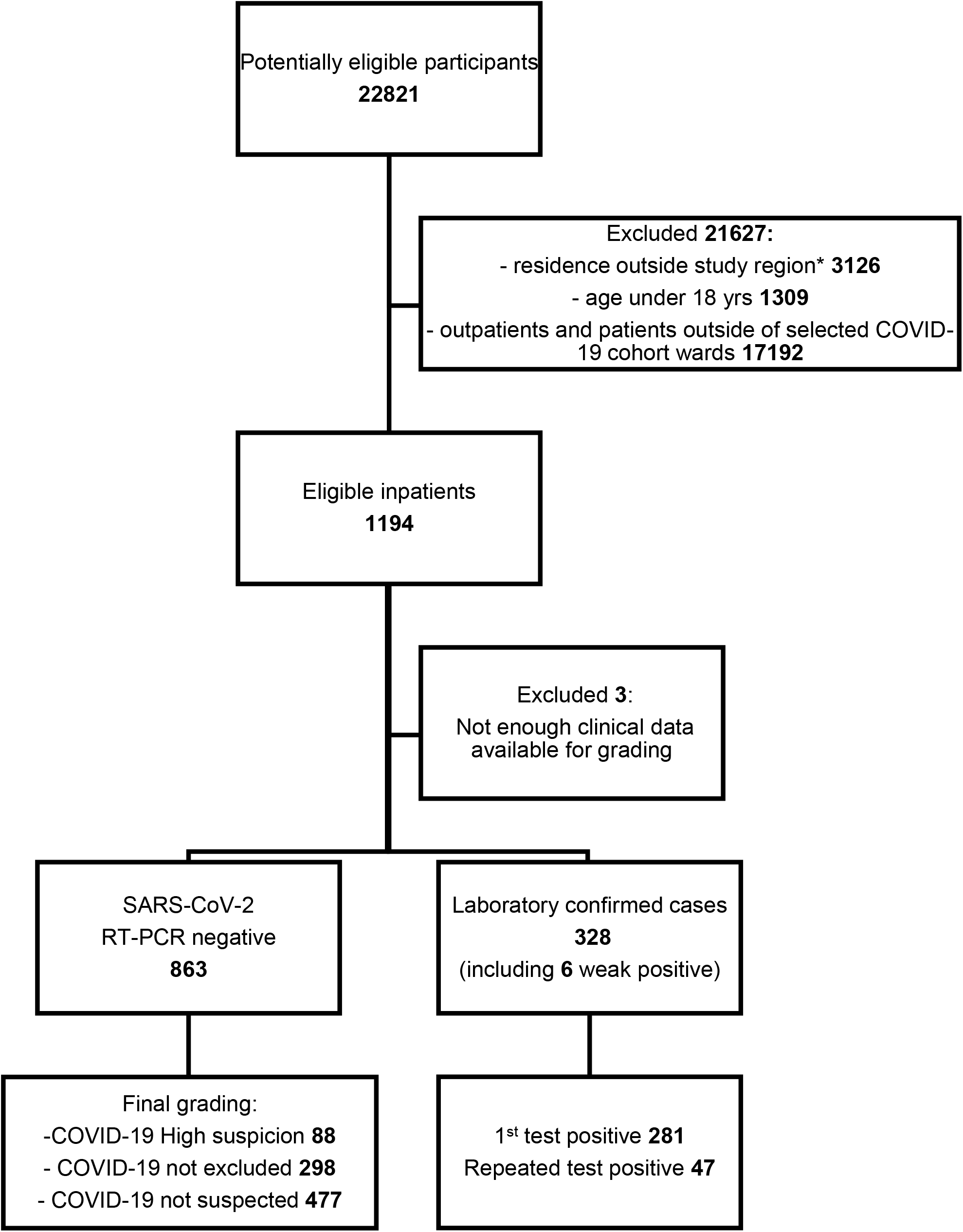
Selection of the inpatient cohort presented as a flowchart.

The descriptive statistics of the tested individuals and subgroups are presented in Table 1, Table 2 and eFigure 2.

**Table 1.**
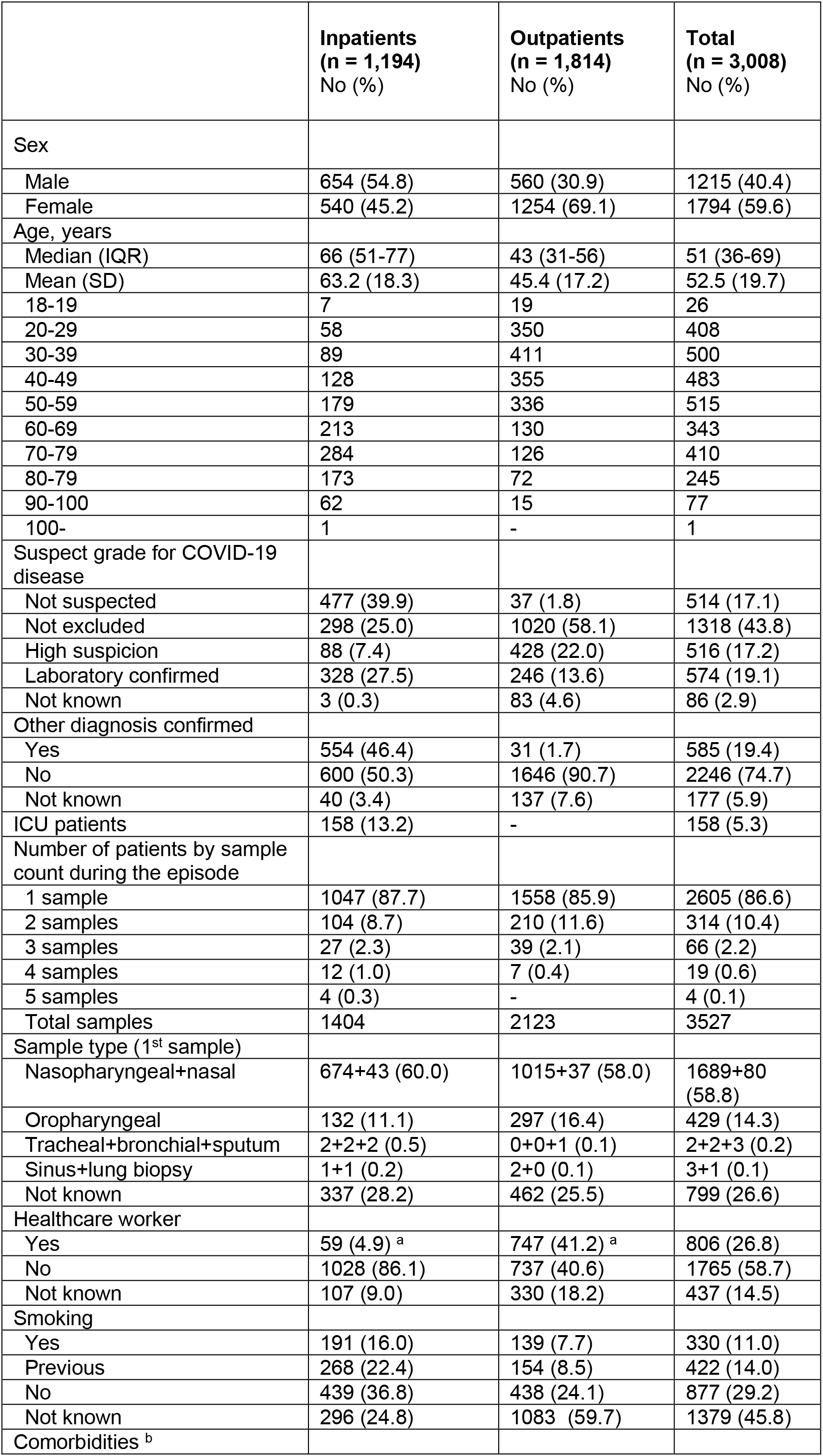
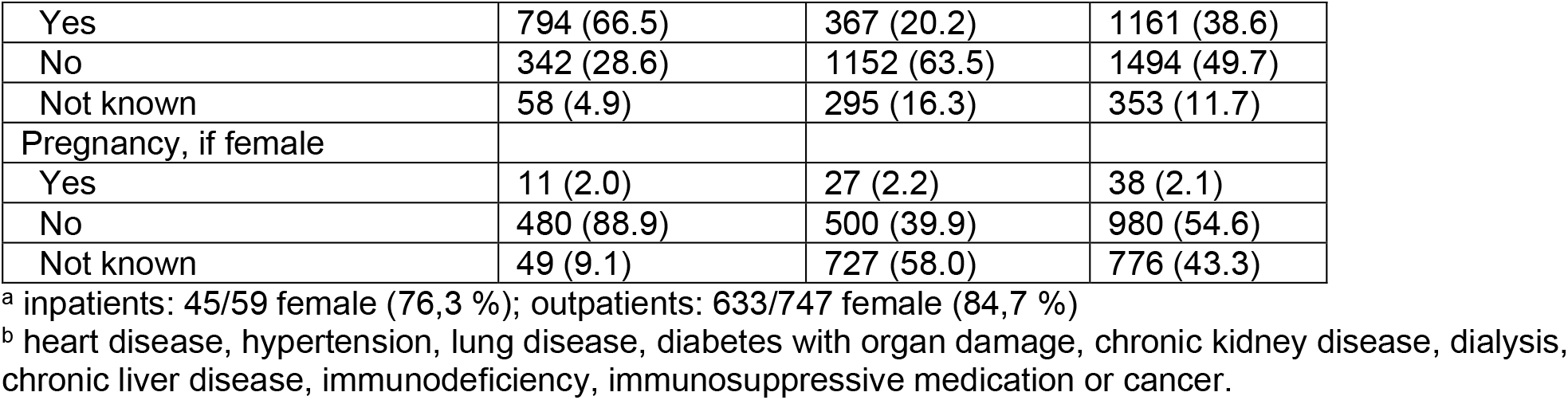
Patient demographic and clinical characteristics.

|  | **Inpatients**<br>**(n = 1,194)**<br>**No (%)** | **Outpatients**<br>**(n = 1,814)**<br>**No (%)** | **Total**<br>**(n = 3,008)**<br>**No (%)** |
| --- | --- | --- | --- |
| Sex |  |  |  |
| Male | 654 (54.8) | 560 (30.9) | 1215 (40.4) |
| Female | 540 (45.2) | 1254 (69.1) | 1794 (59.6) |
| Age, years |  |  |  |
| Median (IQR) | 66 (51-77) | 43 (31-56) | 51 (36-69) |
| Mean (SD) | 63.2 (18.3) | 45.4 (17.2) | 52.5 (19.7) |
| 18-19 | 7 | 19 | 26 |
| 20-29 | 58 | 350 | 408 |
| 30-39 | 89 | 411 | 500 |
| 40-49 | 128 | 355 | 483 |
| 50-59 | 179 | 336 | 515 |
| 60-69 | 213 | 130 | 343 |
| 70-79 | 284 | 126 | 410 |
| 80-79 | 173 | 72 | 245 |
| 90-100 | 62 | 15 | 77 |
| 100- | 1 | - | 1 |
| Suspect grade for COVID-19 disease |  |  |  |
| Not suspected | 477 (39.9) | 37 (1.8) | 514 (17.1) |
| Not excluded | 298 (25.0) | 1020 (58.1) | 1318 (43.8) |
| High suspicion | 88 (7.4) | 428 (22.0) | 516 (17.2) |
| Laboratory confirmed | 328 (27.5) | 246 (13.6) | 574 (19.1) |
| Not known | 3 (0.3) | 83 (4.6) | 86 (2.9) |
| Other diagnosis confirmed |  |  |  |
| Yes | 554 (46.4) | 31 (1.7) | 585 (19.4) |
| No | 600 (50.3) | 1646 (90.7) | 2246 (74.7) |
| Not known | 40 (3.4) | 137 (7.6) | 177 (5.9) |
| ICU patients | 158 (13.2) | - | 158 (5.3) |
| Number of patients by sample count during the episode |  |  |  |
| 1 sample | 1047 (87.7) | 1558 (85.9) | 2605 (86.6) |
| 2 samples | 104 (8.7) | 210 (11.6) | 314 (10.4) |
| 3 samples | 27 (2.3) | 39 (2.1) | 66 (2.2) |
| 4 samples | 12 (1.0) | 7 (0.4) | 19 (0.6) |
| 5 samples | 4 (0.3) | - | 4 (0.1) |
| Total samples | 1404 | 2123 | 3527 |
| Sample type (1<sup>st</sup> sample) |  |  |  |
| Nasopharyngeal+nasal | 674+43 (60.0) | 1015+37 (58.0) | 1689+80 (58.8) |
| Oropharyngeal | 132 (11.1) | 297 (16.4) | 429 (14.3) |
| Tracheal+bronchial+sputum | 2+2+2 (0.5) | 0+0+1 (0.1) | 2+2+3 (0.2) |
| Sinus+lung biopsy | 1+1 (0.2) | 2+0 (0.1) | 3+1 (0.1) |
| Not known | 337 (28.2) | 462 (25.5) | 799 (26.6) |
| Healthcare worker |  |  |  |
| Yes | 59 (4.9) <sup>a</sup> | 747 (41.2) <sup>a</sup> | 806 (26.8) |
| No | 1028 (86.1) | 737 (40.6) | 1765 (58.7) |
| Not known | 107 (9.0) | 330 (18.2) | 437 (14.5) |
| Smoking |  |  |  |
| Yes | 191 (16.0) | 139 (7.7) | 330 (11.0) |
| Previous | 268 (22.4) | 154 (8.5) | 422 (14.0) |
| No | 439 (36.8) | 438 (24.1) | 877 (29.2) |
| Not known | 296 (24.8) | 1083 (59.7) | 1379 (45.8) |
| Comorbidities <sup>b</sup> |  |  |  |
| Yes | 794 (66.5) | 367 (20.2) | 1161 (38.6) |
| No | 342 (28.6) | 1152 (63.5) | 1494 (49.7) |
| Not known | 58 (4.9) | 295 (16.3) | 353 (11.7) |
| Pregnancy, if female |  |  |  |
| Yes | 11 (2.0) | 27 (2.2) | 38 (2.1) |
| No | 480 (88.9) | 500 (39.9) | 980 (54.6) |
| Not known | 49 (9.1) | 727 (58.0) | 776 (43.3) |

<sup>a</sup> inpatients: 45/59 female (76,3 %); outpatients: 633/747 female (84,7 %)
<sup>b</sup> heart disease, hypertension, lung disease, diabetes with organ damage, chronic kidney disease, dialysis, chronic liver disease, immunodeficiency, immunosuppressive medication or cancer.

**Table 2.** Patient demographic and clinical characteristics by clinical suspicion of COVID-19 in inpatients (A) and outpatients (B). Three patients in the inpatient cohort could not be classified. ‘Not suspected’: deemed by the clinician as non-compatible with COVID-19 disease or were diagnosed with another acute disease; ‘Not excluded’: no other diagnosis recorded explaining their current symptoms, and COVID-19 disease could not be excluded. ‘High suspicion’: physician in charge of the treatment recorded the suspicion on clinical grounds to the electronic patient record, OR the patient fulfilled a set of pre-defined clinical and exposure criteria (see Methods); ‘Laboratory confirmed’: tested positive from the first sample taken or with repeated testing with SARS-CoV-2 RT-PCR.

**A**
|  | COVID-19<br>No Suspicion<br>(n=477)<br>Number (%) | COVID-19<br>Not Excluded<br>(n=298)<br>Number (%) | COVID-19<br>High<br>Suspicion<br>(n=88)<br>Number<br>(%) | COVID-19<br>Laboratory<br>Confirmed<br>(n=328)<br>Number (%) | P Value<br>COVID-19<br>High Suspicion<br>vs. Covid<br>lab.confirmed |
| --- | --- | --- | --- | --- | --- |
| Sex |  |  |  |  | .16<sup>a</sup> |
| Male | 255 (53.5) | 172 (57.7) | 41 (46.6) | 183 (55.8) |  |
| Female | 222 (46.5) | 126 (42.3) | 47 (53.4) | 145 (44.2) |  |
| Age, years |  |  |  |  |  |
| Median (IQR) | 71 (56-80) | 69 (52-78) | 55.5 (41-70.25) | 58 (47.75-72) |  |
| Mean (SD) |  |  |  |  |  |
| Range | 18-103 | 18-96 | 18-96 |  |  |
| Other diagnosis confirmed |  |  |  |  |  |
| Yes | 448 | 92 (30.9) | ND | ND |  |
| No | 17 | 184 (61.7) | ND | ND |  |
| Not known | 12 | 22 (7.4) | ND | ND |  |
| Influenza A + Influenza B + RSV |  |  |  |  |  |
| Total tested | 395 | 261 | 79 | 186 | <.001<sup>a</sup> |
| Positive | 6+4+24<br>(1.3+0.8+3.1) | 0+0+9<br>(0+0+3.0) | 0+0+0 | 0+0+0 | NA |
| Other virus finding |  |  |  |  |  |
| Total tested | 15 | 20 | 7 | 1 | <.001<sup>a</sup> |
| Coronavirus<br>229E/NL63/OC43 | 2 |  | - | - | NA |
| Rhinovirus | - | 1 | - | - | NA |
| Rhinovirus and<br>Coronavirus<br>229E/NL63/OC43 | 1 |  | - | - | NA |
| ICU patients | 44 (9.2) | 22 (7.4) | 7 (7.8) | 82 (25.0) | <.001<sup>a</sup> |
| Time between symptom<br>onset and first SARS-CoV-2 RT-PCR test |  |  |  |  |  |
| <1 day | 152 (31.9) | 71 (23.8) | 7 (8.0) | 22 (6.7) |  |
| 1-2 days | 112 (23.5) | 59 (19.8) | 12 (13.6) | 55 (16.8) |  |
| 3-4 days | 46 (9.6) | 39 (13.1) | 15 (17.0) | 50 (15.2) |  |
| 5-6 days | 29 (6.1) | 28 (9.4) | 7 (8.0) | 58 (17.7) |  |
| 7-14 days | 61 (12.8) | 54 (18.1) | 38 (43.2) | 132 (40.2) |  |
| >14 days | 38 (8.0) | 31 (10.4) | 7 (8.0) | 8 (2.4) |  |
| No data | 39 (8.2) | 16 (5.4) | 2 (2.3) | 3 (0.9) |  |
| Fever |  |  |  |  | <.001<sup>b</sup> |
| Yes | 233 (48.8) | 193 (64.8) | 67 (76.1) | 300 (91.5) |  |
| No | 187 (39.2) | 85 (28.5) | 20 (22.7) | 21 (6.4) |  |
| Not known | 57 (11.9) | 20 (6.7) | 1 (1.1) | 7 (2.1) |  |
| Upper respiratory<br>symptoms |  |  |  |  | .86<sup>a</sup> |
| Yes | 119 (24.9) | 83 (27.9) | 38 (43.2) | 131 (39.9) |  |
| No | 218 (45.7) | 109 (36.6) | 23 (26.1) | 90 (27.4) |  |
| Not known | 140 (29.4) | 106 (35.6) | 27 (30.7) | 107 (32.6) |  |
| Lower respiratory symptoms |  |  |  |  | .52<sup>b</sup> |
| Yes | 282 (59.1) | 240 (80.5) | 84 (95.5) | 299 (91.2) |  |
| No | 134 (28.1) | 39 (13.1) | 2 (2.3) | 14 (4.3) |  |
| Not known | 61 (12.8) | 19 (6.4) | 2 (2.3) | 15 (4.6) |  |
| Infectious finding in chest X-ray/CT |  |  |  |  | .33<sup>b</sup> |
| Yes | 78 (16.4) | 103 (34.6) | 72 (81.8) | 266 (81.1) |  |
| No | 337 (70.6) | 181 (60.7) | 12 (13.6) | 55 (16.8) |  |
| Not done | 28 (5.9) | 9 (3.0) | 2 (2.3) | 2 (0.6) |  |
| Not known | 34 (7.1) | 5 (1.7) | 2 (2.3) | 5 (1.5) |  |
| Altered sense of smell |  |  |  |  | .24<sup>a</sup> |
| Yes | 3 (0.6) | 2 (0.7) | 3 (3.4) | 27 (8.2) |  |
| No | 59 (12.4) | 24 (8.1) | 13 (14.8) | 38 (11.6) |  |
| Not known | 415 (87.0) | 272 (91.3) | 72 (81.8) | 263 (80.2) |  |
| Gastrointestinal symptoms |  |  |  |  | .03<sup>a</sup> |
| Yes | 137 (28.7) | 65 (21.8) | 32 (36.4) | 155 (47.3) |  |
| No | 146 (30.6) | 109 (36.6) | 21 (23.9) | 90 (27.4) |  |
| Not known | 194 (40.7) | 124 (41.6) | 35 (39.8) | 83 (25.3) |  |
| Thrombosis |  |  |  |  | .26<sup>b</sup> |
| Yes | 18 (3.8) | 9 (3.0) | 6 (6.8) | 10 (3.0) |  |
| No | 261 (54.7) | 208 (69.8) | 56 (63.6) | 220 (67.1) |  |
| Not known | 198 (41.5) | 81 (27.2) | 26 (29.5) | 98 (29.9) |  |
| Contact in 14 days to symptomatic COVID-19 |  |  |  |  | .03<sup>a</sup> |
| Yes | 15 (3.1) | 10 (3.4) | 18 (20.5) | 117 (35.7) |  |
| No | 214 (44.9) | 124 (41.6) | 33 (37.5) | 101 (30.8) |  |
| Not known | 248 (52.0) | 164 (55.0) | 37 (42.0) | 110 (33.5) |  |
| Travel in 14 days |  |  |  |  | .11<sup>a</sup> |
| To defined risk areas <sup>c</sup> | 7 (1.5) | 3 (1.0) | 4 (4.5) | 28 (8.5) |  |
| Other | 11 (2.3) | 12 (4.0) | 10 (11.4) | 19 (5.8) |  |
| No travel abroad | 300 (62.9) | 208 (69.8) | 55 (62.5) | 227 (69.2) |  |
| Not known | 159 (33.3) | 75 (25.2) | 19 (21.6) | 54 (16.5) |  |
| Healthcare worker: |  |  |  |  | .13<sup>b</sup> |
| Yes | 10 (2.1) | 8 (2.7) | 5 (5.7) | 36 (11.0) |  |
| No | 432 (90.6) | 269 (90.3) | 68 (77.3) | 256 (78.0) |  |
| Not known | 35 (7.3) | 21 (7.0) | 15 (17.0) | 36 (11.0) |  |
| Clinical severity grade |  |  |  |  | .23<sup>a</sup> |
| Low | ND | ND | - | 2 (0.6) |  |
| Medium | ND | ND | 62 (70.5) | 198 (60.4) |  |
| Severe <sup>d</sup> | ND | ND | 23 (26.1) | 120 (36.6) |  |
| Not known | ND | ND | 3 (3.4) | 8 (2.4) |  |
| Smoking |  |  |  |  | .001<sup>a</sup> |
| Yes | 106 (22.2) | 57 (19.1) | 13 (14.8) | 14 (4.3) |  |
| Quitted | 95 (19.9) | 87 (29.2) | 19 (21.6) | 66 (20.1) |  |
| No | 137 (28.7) | 88 (29.5) | 37 (42.0) | 177 (54.0) |  |
| Not known | 139 (29.1) | 66 (22.1) | 19 (21.6) | 71 (21.6) |  |
| Comorbidities |  |  |  |  | .19<sup>b</sup> |
| Yes | 364 (76.3) | 230 (77.2) | 49 (54.4) | 150 (45.7) |  |
| No | 80 (16.8) | 66 (22.1) | 39 (43.3) | 158 (48.2) |  |
| Not known | 33 (6.9) | 2 (0.7) | 2 (2.2) | 20 (6.1) |  |

<sup>a</sup> Chi-squared test
<sup>b</sup> Fisher exact test
<sup>c</sup> defined risk areas were Austria Tirol, Northern Italy, Spain, China and South Korea
<sup>d</sup> admitted to ICU OR recorded respiratory rate of $\geq 30/\text{min}$ OR oxygen saturation $\leq 85\%$ (with or without supplemental oxygen at any stage)
ND = not determined

B
|  | COVID-19<br>No Suspicion<br>(n=37)<br>Number (%) | COVID-19<br>Not Excluded<br>(n=1020)<br>Number (%) | COVID-19<br>High<br>Suspicion<br>(n=428)<br>Number<br>(%) | COVID-19<br>Laboratory<br>Confirmed<br>(n=246)<br>Number<br>(%) | P Value<br>COVID-19<br>High<br>Suspicion vs.<br>Covid<br>lab.confirmed |
| --- | --- | --- | --- | --- | --- |
| Sex |  |  |  |  | .004<sup>a</sup> |
| Male | 17 (45.9) | 256 (25.1) | 151 (35.3) | 115 (46.7) |  |
| Female | 20 (54.1) | 764 (74.9) | 277 (64.7) | 131 (53.3) |  |
| Age, years |  |  |  |  |  |
| Median (IQR) | 61 (34-76) | 45 (32-58) | 40 (30-51) | 42 (31-55) |  |
| Mean (SD) |  |  |  |  |  |
| Range | 18-89 | 18-98 | 18-91 | 18-94 |  |
| Other diagnosis confirmed |  |  |  |  | .13<sup>b</sup> |
| Yes | 29 (78.4) | - | - | - |  |
| No | 7 (18.9) | 957 (93.8) | 417 (97.4) | 234 (95.1) |  |
| Not known | 1 (2.7) | 63 (6.2) | 11 (2.6) | 12 (4.9) |  |
| Influenza A + Influenza B + RSV |  |  |  |  |  |
| Total tested | 21 | 104 | 58 | 17 | .01<sup>a</sup> |
| Positive | 3+4+2 | 0+0+0 | 0+0+0 | 0+0+0 | NA |
| Other virus finding |  |  |  |  |  |
| Total tested | 4 | 2 | 3 | 1 | >.99<sup>b</sup> |
| Rhinovirus | 1 | - | - | - | NA |
| Time between symptom onset<br>and first SARS-CoV-2 RT-PCR<br>test |  |  |  |  |  |
| <1 day | 5 (13.5) | 28 (2.7) | 14 (3.3) | 15 (6.1) |  |
| 1-2 days | 12 (32.4) | 260 (25.5) | 120 (28.0) | 74 (30.1) |  |
| 3-4 days | 4 (10.8) | 203 (19.9) | 84 (19.6) | 52 (21.1) |  |
| 5-6 days | 7 (18.9) | 117 (11.5) | 48 (11.2) | 30 (12.2) |  |
| 7-14 days | 3 (8.1) | 246 (24.1) | 98 (22.9) | 40 (16.3) |  |
| >14 days | 1 (2.7) | 65 (6.4) | 19 (4.4) | 5 (2.0) |  |
| No data | 5 (13.5) | 101 (9.9) | 45 (10.5) | 30 (12.2) |  |
| Fever |  |  |  |  | <.001<sup>a</sup> |
| Yes | 11 (29.7) | 122 (12.0) | 55 (12.9) | 90 (36.6) |  |
| No | 16 (43.2) | 337 (33.0) | 144 (33.6) | 59 (24.0) |  |
| Not known | 10 (27.0) | 561 (55.0) | 229 (53.5) | 97 (39.4) |  |
| Upper respiratory symptoms |  |  |  |  | .81<sup>b</sup> |
| Yes | 10 (27.0) | 371 (36.4) | 179 (41.8) | 102 (41.5) |  |
| No | 7 (18.9) | 15 (1.5) | 6 (1.4) | 5 (2.0) |  |
| Not known | 20 (54.1) | 634 (62.2) | 243 (56.8) | 139 (56.5) |  |
| Lower respiratory symptoms |  |  |  |  | .51<sup>a</sup> |
| Yes | 19 (54.3) | 440 (43.1) | 198 (46.3) | 123 (50.0) |  |
| No | 4 (10.8) | 29 (2.8) | 11 (2.6) | 8 (3.3) |  |
| Not known | 14 (37.8) | 551 (54.0) | 219 (51.2) | 115 (46.7) |  |
| Infectious finding in chest X-ray/CT |  |  |  |  | .18<sup>b</sup> |
| Yes | 1 (2.7) | 4 (0.4) | 4 (0.9) | 5 (2.0) |  |
| No | 14 (37.8) | 57 (5.6) | 9 (2.1) | 11 (4.5) |  |
| Not done | 16 (43.2) | 613 (61.0) | 244 (57.0) | 130 (52.8) |  |
| Not known | 6 (16.2) | 346 (33.9) | 171 (40.0) | 100 (40.7) |  |
| Altered sense of smell |  |  |  |  | <.001<sup>b</sup> |
| Yes | 1 (2.7) | 5 (0.5) | 8 (1.9) | 23 (9.3) |  |
| No | 1 (2.7) | 2 (0.2) | - | 1 (0.4) |  |
| Not known | 35 (94.6) | 1013 (99.3) | 420 (98.1) | 222 (90.2) |  |
| Gastrointestinal symptoms |  |  |  |  | .19<sup>b</sup> |
| Yes | 2 (5.4) | 55 (5.4) | 15 (3.5) | 14 (5.7) |  |
| No | 5 (13.5) | 9 (0.9) | 1 (0.2) | 2 (0.8) |  |
| Not known | 30 (81.1) | 956 (93.7) | 412 (96.3) | 230 (93.5) |  |
| Thrombosis |  |  |  |  | .49<sup>b</sup> |
| Yes | - | - | - | - |  |
| No | 17 (45.9) | 117 (11.5) | 36 (8.4) | 25 (10.2) |  |
| Not known | 20 (54.1) | 903 (88.5) | 392 (91.6) | 221 (89.8) |  |
| Contact in 14 days to symptomatic COVID-19 |  |  |  |  | <.001<sup>a</sup> |
| Yes | 7 (18.9) | 3 (0.3) | 299 (69.9) | 141 (57.3) |  |
| No | 12 (32.4) | 341 (33.4) | 32 (7.5) | 16 (6.5) |  |
| Not known | 18 (48.6) | 676 (66.3) | 97 (22.4) | 89 (36.2) |  |
| Travel in 14 days |  |  |  |  | .005<sup>a</sup> |
| To defined risk areas <sup>c</sup> | 3 (8.1) | 12 (1.2) | 131 (30.6) | 49 (19.9) |  |
| Other | - | 197 (19.3) | 16 (3.7) | 16 (6.5) |  |
| No travel abroad | 24 (64.9) | 626 (61.4) | 205 (47.9) | 121 (49.2) |  |
| Not known | 10 (27.0) | 185 (18.1) | 76 (17.8) | 60 (24.4) |  |
| Healthcare worker: |  |  |  |  | <.001<sup>a</sup> |
| Yes | 6 (16.2) | 533 (52.3) | 118 (27.6) | 59 (24.0) |  |
| No | 28 (75.7) | 398 (39.0) | 170 (39.7) | 127 (51.6) |  |
| Not known | 3 (8.1) | 89 (8.7) | 140 (32.9) | 60 (24.4) |  |
| Clinical severity grade: |  |  |  |  | .001<sup>b</sup> |
| Low | ND | ND | 423 (98.8) | 229 (93.1) |  |
| Medium | ND | ND | 2 (0.5) | 2 (0.8) |  |
| Severe | ND | ND | - | - |  |
| Not known | ND | ND | 3 (0.7) | 15 (6.1) |  |
| Smoking: |  |  |  |  | .80<sup>b</sup> |
| Yes | 3 (8.1) | 92 (9.0) | 29 (6.8) | 12 (4.9) |  |
| Quitted | 8 (21.6) | 107 (10.5) | 24 (5.6) | 14 (5.7) |  |
| No | 9 (24.3) | 270 (26.5) | 95 (22.2) | 55 (22.4) |  |
| Not known | 17 (45.9) | 551 (54.0) | 280 (65.4) | 165 (67.1) |  |
| Comorbidities: |  |  |  |  | >.99<sup>a</sup> |
| Yes | 19 (51.4) | 248 (24.3) | 56 (13.1) | 33 (13.4) |  |
| No | 15 (40.5) | 653 (64.0) | 285 (66.6) | 163 (66.3) |  |
| Not known | 3 (8.1) | 119 (11.7) | 87 (20.3) | 50 (20.3) |  |

<sup>a</sup> Chi-squared test
<sup>b</sup> Fisher exact test
<sup>c</sup> defined risk areas were Austria Tiroli, Northern Italy, Spain, China and South Korea
ND = not determined
NA = not applicable

### Index testing

SARS-CoV-2 RT-PCR testing was conducted by one of the following methods: Cobas® SARS-CoV-2 test kit on the CobasC® 6800 system (Roche Diagnostics, Basel, Switzerland), Amplidiag® COVID-19 test (Mobidiag, Espoo, Finland,) and a laboratory-developed test based on a protocol recommended by WHO.^16^ The specifics and analytical performance of these methods in our laboratory setting have been described previously.^4^ Samples were collected with nasopharyngeal swabs (FLOQSwab, Copan, Brescia, Italy) but oropharyngeal swabs were used in a proportion of patients due to global shortage of nasopharyngeal swabs.

Samples were analysed in median 24 hours after collection. Samples with failed results were retested and only qualified results were included. 13 weak positive results which became positive in >35 PCR cycles (7 outpatients and 6 inpatients) were included.

### Reference standard used in the study

Since no gold standard for COVID-19 diagnosis exists, we decided to use high clinical suspicion for COVID-19 as the reference standard for the RT-PCR test. We systemically graded the clinical suspicion for COVID-19 based on a combination of symptoms, clinical findings, and recorded exposure to laboratory confirmed COVID-19 cases or travel history to epidemic areas. The criteria were based on CDC’s and ECDC’s case definitions for COVID-19 in April 2020. Electronic patient records or line lists were reviewed by a team consisting of senior residents in Infectious Diseases and Clinical Microbiology, medical students, and research nurses. Patients’ medical history, symptoms, and epidemiological information were collected into a Microsoft Access® database according to the pre-defined criteria. Chest X-ray and CT findings indicating chest infection were recorded according to radiologist’s interpretation. The team collecting the data were aware of the SARS-CoV-2 RT-PCR test result when collecting the data.

The clinical suspicion for COVID-19 disease was graded as follows:

1. ‘Not suspected’ patients were deemed by the clinician as non-compatible with COVID-19 disease or were diagnosed with another acute disease.
2. ‘Not excluded’ patients had no other diagnosis recorded explaining their current symptoms, and COVID-19 disease could not be excluded.
3. ‘High suspicion’ patients were considered to suffer from a probable COVID-19 if the physician in charge of the treatment recorded the suspicion on clinical grounds to the electronic patient record, OR the patient fulfilled at least one of the following criteria:
  a. respiratory symptoms and/or fever and/or diagnostic finding for infection in chest X-ray/CT and travel history to epidemic regions at the time of the study i.e. Tirol/Austria, Northern Italy, Spain, Iran, South Korea, or China during the preceding 14 days.
  b. respiratory symptoms and fever and diagnostic finding in chest X-ray/CT during April 2020 (time criterion based on the changed epidemiological situation).
  c. respiratory or gastrointestinal symptoms or fever or diagnostic finding in chest X-ray/CT and a close contact with a laboratory confirmed COVID-19 patient during the preceding 14 days prior to disease onset.
4. ‘Laboratory confirmed’ patients were tested positive for SARS-CoV-2 RT-PCR during the study period.

### Sample size calculation

We estimated the minimum sample size needed for outpatients based on Bujang et al^17^, with a minimal statistical power of 80% and type I error <0.05. Sample size calculation for sensitivity requires a prevalence estimation in the target population. During the study period, the median positivity rate was 9.6% (eFigure 1) so we estimated a 10% prevalence for the tested population. Published estimates from small cohorts^18-19^ available at the time reported sensitivity of the SARS-CoV-2 RT-PCR to be on average 70%. Based on these estimates the minimum sample size of outpatients for null hypothesis of sensitivity of 70% was 1,550. We performed another sample size calculation by using the nomogram described by Carley et al.^20^ which accounts for confidence intervals (CI): with CI of 93% and prevalence 10%, 70% sensitivity would require a minimum sample size of 1,600.

### Group comparisons

To detect if the high suspicion and the laboratory confirmed groups were comparable, we compared demographic and clinical characteristics between them. To compare these two groups with respect to the categorical variables, we used the Pearson’s Chi-squared test without or with Yate’s correction for continuity or the Fisher’s exact test, as appropriate. For the extensive contingency tables with the excess of small (expected) frequencies, we assessed the simulated p-value of the Fisher’s test based on 20,000 replicates. The differences in the age distribution were assessed using the Mann-Whitney U-test. These comparisons were performed separately within the inpatients and outpatients.

### Analysis of sensitivity

For the laboratory confirmed patients, the sensitivity values were calculated based on the first RT-PCR test of each patient. All patients who tested RT-PCR positive during a specific disease episode were considered laboratory confirmed. Of these, the first samples with a negative RT-PCR test result were considered false negatives, while the first samples with a positive result were considered true positives. The same disease episode would include samples taken ≤14 days apart.

The 95% CIs for (binomial) sensitivity were calculated by using the Wilson-Score method, which is based on inverting the z-test for a single proportion and provides more reliable coverage than the alternatives. We performed comparisons of sensitivity between the subgroups by using the independent sample tests for binomial proportions, including Chi-squared test without or with Yate’s correction for continuity or the Fisher’s exact test, as appropriate. For the extensive contingency tables with the excess of small (expected) frequencies, we assessed the simulated p-value of the Fisher’s test based on 20,000 replicates. We set the confidence level at 5%. All calculations were performed using the R software.

## Results

### Demographics of the study population and clinical comparison between study groups

In all, 3,008 individuals were eligible for this study (Figure 1): 1,814 outpatients and 1,194 inpatients. Altogether 83 eligible outpatients (4.6%) and 3 inpatients (0.3%) were excluded from the final analysis due to insufficient data for the grading of clinical suspicion.

The inpatients were on average older than outpatients, comorbidities were more common, and the male sex was slightly overrepresented (Table 1). Healthcare workers and women were overrepresented in the outpatient population reflecting the distribution of the whole tested population, as reported before.^21^

All patients were categorized by a clinical grade of suspicion for COVID-19 as described in Methods (Table 1). To detect if our grading created systematic bias, we compared test negative patients that were deemed as high suspicion to laboratory confirmed patients. There were no significant differences in sex or age distribution between these groups, but patients treated in the intensive care unit were overrepresented in the laboratory confirmed patients (Table 2). Laboratory confirmed patients were also more often febrile and had had contact with laboratory confirmed symptomatic COVID-19 cases.

In the outpatients, the high suspicion group had a higher proportion of females and healthcare workers compared to laboratory confirmed cases. This was expected based on the overall higher testing rate of both.^21^ Again, the laboratory confirmed cases were more often febrile. Since our grading criteria included exposure to symptomatic COVID-19 patients or travel to epidemic areas, these factors were more common in the high suspicion group that tested negative, than in the laboratory confirmed group (Table 2).

### Sensitivity of the first SARS-CoV-2 RT-PCR in inpatients and outpatients

The sensitivity of SARS-CoV-2 RT-PCR was calculated with two different denominators (Table 3). We first calculated the sensitivity with laboratory confirmed cases, i.e. repeat-tested patients as a denominator, yielding the highest sensitivity estimates in this study, as follows: 85.7% for inpatients; 95.5% for outpatients, and 89.9% for all. Due to low number of repeat-tested patients (N=11), the calculation for outpatients here is unreliable.

**Table 3.** Sensitivity calculations for the first SARS-CoV-2 RT-PCR test. The numerator for all calculations is the number of patients that tested positive with first sample.

| **Denominator** |  | **Inpatients** | **Outpatients** | **All** |
| --- | --- | --- | --- | --- |
| COVID-19 Laboratory confirmed patients | N/N | 281/328 | 235/246 | 516/574 |
|  | **Sensitivity (%)** | **85.7%** | **95.5%** | **89.9%** |
|  | 95% CI | (81.5–89.1%) | (92.2–97.5%) | (88.2–92.1%) |
| COVID-19 Laboratory confirmed + High suspicion patients | N/N | 281/416 | 235/674 | 516/1090 |
|  | **Sensitivity (%)** | **67.5%** | **34.9%** | **47.3%** |
|  | 95% CI | (62.9–71.9%) | (31.4–38.5%) | (44.4–50.3%) |

The sensitivity was then calculated by including in the denominator patients that were graded as high suspicion but never tested positive, yielding the following sensitivity values: 67.5% for inpatients, 34.9% for outpatients and 47.3% for all. Thus, the lowest calculated sensitivity estimate in this study was for outpatients with high suspicion.

The delay between disease onset and testing was longer for inpatients than outpatients (Table 2). We could not detect a significant difference in the delay to first test between the laboratory confirmed cases and the high suspicion group in either cohort (eFigure 3, Fisher’s Exact Test p=1 when “No data” category excluded). However, for inpatients, information on the delay was missing more often in the high suspicion group (2.3%) compared to the laboratory confirmed (0.9%, p=0.026) (Table 2). For outpatients, information on the delay was missing less often in the high suspicion group (10.5%) as compared to the laboratory confirmed (12.2% p=0.026) (Table 2).

We could not detect a significant difference between the sensitivity of nasopharyngeal and oropharyngeal samples in the inpatients (p= .51, Chi-squared test), outpatients (p= .22) or all patients (p= .66) (eTable 3; eFigure 4). However, data on the specimen type was missing in 20.4% (inpatients) and 17.4% (outpatients) of the cases.

### Delay between symptom onset and positive test result

To estimate the delay from disease onset for highest clinical sensitivity for SARS-CoV-2 RT-PCR, we calculated sensitivities for different time frames. To achieve reliable group sizes, both cohorts were pooled together. There was no significant difference in the test sensitivity according to delay from onset, calculated for the laboratory confirmed cases alone, and with the high clinical suspicion group included (P=0.1013 Fisher’s Exact Test for Count Data with simulated p-value, based on 20000 replicates; Figure 2). Detailed sensitivity calculations per delay from disease onset are presented in eTable 4.

**Figure 2.**
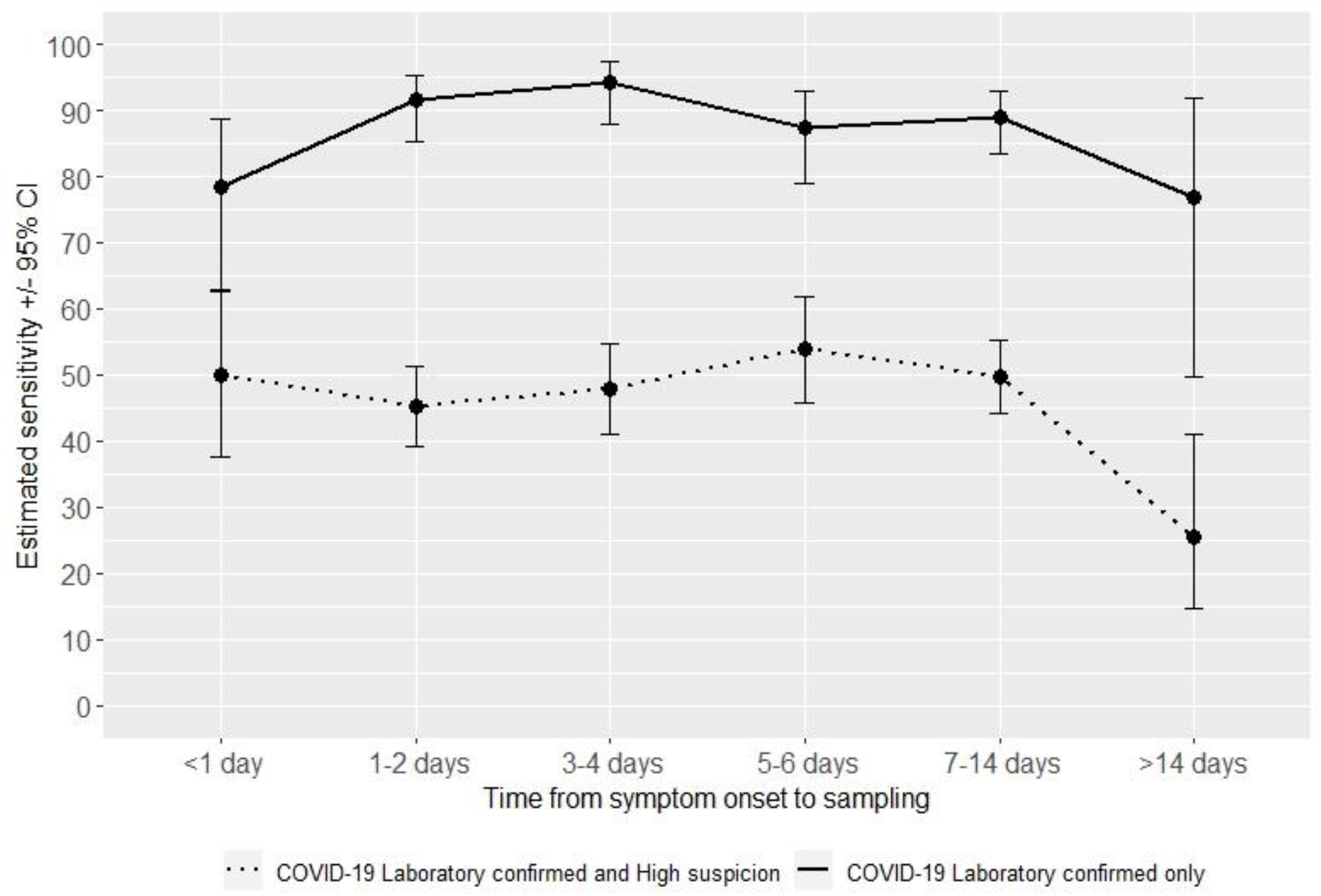
Clinical SARS-CoV-2 RT-PCR sensitivity estimates in the laboratory confirmed, and in the laboratory confirmed and high suspicion group combined presented according to delay (days) from symptom onset to sampling.

## Discussion

Wide-spread testing and contact tracing together with social distancing has been promoted as the tool that prevents new lockdowns – without clear understanding of how well the SARS-CoV-2 RT-PCR test performs. Here, we used clinical suspicion as the gold standard to estimate the clinical sensitivity of the test.

A previous large-scale sensitivity estimate for SARS-CoV-2 molecular testing was based on repeat-tested individuals.^9,22^ This approach overestimates the sensitivity. Repeated testing is done mostly on inpatients who have a strong clinical suspicion, rendering high pre-test probability. We sought to overcome this limitation by including a large cohort of outpatients. From an epidemiological point-of-view, understanding the clinical sensitivity for mild cases is important. A recent preprint reported an RT-PCR sensitivity of 64% for exposed family members systemically tested with serology.^23^ This is in line with our sensitivity estimation for inpatients.

Our analysis was done in a low prevalence setting. Thus, the negative predictive value for the RT-PCR test was high (89%) for the outpatients even though the clinical sensitivity was low (35%), assuming all COVID-19 excluded cases were true negatives. In light of clinical judgement, false negative rate was high which could reduce the negative predictive value of testing. This is particularly problematic when the prevalence of the disease increases. In such settings, it will impair effective use of wide-spread testing.

For health-care facilities the message of our data is different: a single negative result cannot be trusted to rule out COVID-19 in patients with suitable symptoms. Our data show that the sensitivity of the repeat-tested inpatients was high (86%), and in line with previous reports on repeated testing.^9,22^ When the sensitivity of the COVID-19 PCR test was judged based on the laboratory confirmed and high clinical suspicion patients the estimated sensitivity of the test dropped to around 68%. Our results emphasize the importance of repeated sampling but it also highlights the importance to evaluate the patient’s clinical presentation carefully.

This study estimated test sensitivity both with repeat-tested patients and by using clinical suspicion as a gold standard. The estimated sensitivity (89.9%) for repeat-tested patients is probably an overestimation. In contrast, the estimate which included both laboratory confirmed and high suspicion outpatients (34.9%), is likely an underestimation as COVID-19 symptoms are shared with other respiratory infections, although circulation of e.g. influenza was very limited at the time. The group which included both laboratory confirmed and high suspicion inpatients likely yielded the most realistic sensitivity estimate (67.5%).

Generally the first symptomatic days are considered best for virus detection from the upper airways.^24-25^ Due to the limited sample size our analysis could not detect a definitive time-point for highest sensitivity. However, even with this under-powered estimation, we should have detected major trends.

The study had several limitations. All patients were considered symptomatic so the estimates cannot be generalized to asymptomatic patients. The clinical criteria were set based on the information available in April 2020. While the core symptoms have remained the same, understanding of COVID-19 presentations has since increased. Potential information bias was introduced by the sometimes undetailed clinical records of outpatients. Reporting bias for more detailed symptoms most likely exists, especially for the laboratory confirmed outpatient cases. The specimen types recorded in the sample referrals may have contained errors. While pre-defined clinical criteria were used for grading, the data were collected retrospectively and the data collectors were aware of the index test result.

Large scale molecular testing has permanently changed the practice of clinical microbiology. RT-PCR for SARS-CoV-2 detection has many limitations as a labor intensive test with a relatively slow throughput. This has led to unbearable delays in results. Multiple solutions are being developed: point-of-care viral antigen detection,^26^ sample pooling,^27^ and self-sampling.^28^ All these approaches, which use RT-PCR as a reference, quite consistently report lower sensitivity than RT-PCR. It is thus evident that all our current testing options are far from optimal in detecting all COVID-19 cases. In controlling of the ongoing pandemic, we need focused research to find an appropriate balance in the tradeoff between test sensitivity, and speed and ease of testing in each epidemiological setting.

## Supporting information

eTable OR eFigure

## Data Availability

Researchers may contact study principal investigators to request access to data.

## Author Contributions

Drs Kekäläinen and Kurkela had full access to all of the data in the study and take responsibility for the integrity of the data and the accuracy of the data analysis. Drs Kekäläinen and Kurkela contributed equally to this article. In addition, Dr. Kortela, Mr. Kirjavainen, Dr. Ahava, and Ms. Jokiranta contibuted equally to this article.

## Concept and design

Kortela, Kirjavainen, Ahava, But, Jääskeläinen AE, Lappalainen, Kreivi, Jarva, Kurkela, Kekäläinen

## Acquisition, analysis, or interpretation of data

Kortela, Kirjavainen, Ahava, Jokiranta, But, Lindahl, Lappalainen, Kreivi, Jarva, Kurkela, Kekäläinen

## Drafting of the manuscript

Kekäläinen, Kurkela

## Critical revision of the manuscript for important intellectual content

Kortela, Kirjavainen, Ahava, Jokiranta, But, Lindahl, Jääskeläinen AE, Jääskeläinen AJ, Järvinen, Jokela, Kallio-Kokko, Loginov, Mannonen, Ruotsalainen, Sironen, Vapalahti, Lappalainen, Kreivi, Jarva, Kurkela, Kekäläinen

## Statistical analysis

But, Kirjavainen, Kekäläinen

## Obtained funding

Kekäläinen, Lappalainen

## Administrative, technical, or material support

Jääskeläinen AE, Jääskeläinen AJ, Järvinen, Jokela, Kallio-Kokko, Loginov, Mannonen, Ruotsalainen, Sironen, Vapalahti

## Supervision

Kekäläinen, Kurkela, Jarva, Lappalainen

## Additional Contributions

We thank study nurses Kerstin Ahlskog and Kirsi Lindgren (Dept. of Respiratory Medicine, Helsinki University Hospital) for their help in data collection, and Dr. Lasse Lönnqvist and Prof. Marjukka Myllärniemi for their support during the study and valuable comments on the manuscript.

