## Supplementary material for "Real-life clinical sensitivity of SARS-CoV-2 RT-PCR test in symptomatic patients": eTable OR eFigure

**eTable 1**. List of SARS-CoV-2 RT-PCR testing criteria over the study period.

**eTable 2.** List of COVID-19 cohort wards. Inpatients were admitted to these COVID-19 cohort wards during the study period. Both laboratory confirmed and suspected COVID-19 patients were admitted to the cohort wards, except for two wards (COV_KNKINF and COV_KITEHO) to which only laboratory confirmed cases were admitted.

**eTable 3**. Estimated SARS-CoV-2 RT-PCR sensitivity values in the laboratory confirmed and high suspicion group combined according to specimen type in the first SARS-CoV-2 RT-PCR test.

**eTable 4.** Estimated SARS-CoV-2 RT-PCR sensitivity values in the laboratory confirmed, and in the laboratory confirmed and high suspicion group combined according to delay (days) from symptom onset to first SARS-CoV-2 RT-PCR test.

**eFigure 1.** Number of SARS-CoV-2 RT-PCR tests conducted at the HUSLAB Clinical Microbiology laboratory, and positivity rate over the study period 4 March – 15 April 2020. Median and mean positivity rates were 9.6% and 10%, respectively.

**eFigure 2**. Age distribution of the study population as histograms. A. Inpatients. B. Outpatients.

**eFigure 3** Delay to first SARS-CoV2 RT-PCR test (days) in the laboratory confirmed cases and the high suspicion group. A. Inpatients B. Outpatients.

**eFigure 4** Visualisation of clinical sensitivity estimates for inpatients and outpatients according to specimen type.

**eTable 1**. List of SARS-CoV-2 RT-PCR testing criteria over the study period.

| Time period | SARS-CoV-2 testing criteria | Epidemic areas |
|---|---|---|
| March 6th - March 9th 2020 | 1) Clinical picture of an acute respiratory infection: e.g. fever and/or cough and/or breathing difficulty and/or pneumonic imaging finding<br><br>AND 2) Possible exposure: travel to epidemic area or contact with laboratory confirmed<br><br>OR 3) Admission to ICU for respiratory infection, causative agent unknown | Mainland China, South Korea, Iran, Italy |
| March 9th - March 15th 2020 | Same as 6 March with an expanded epidemic area | Mainland China, South Korea, Iran, Italy, *Tyrol, Nordrhein-Westfalen (Germany)*[*] |
| March 15th - March 22nd 2020 | 1) Quarantined individual experiencing symptoms of respiratory infection;<br><br>2) Admission to hospital for respiratory infection, causative agent unknown;<br><br>3) Individual over the age of 70 presenting with symptoms of respiratory infection;<br><br>4) Symptoms of respiratory infection AND possible exposure: travel or contact with lab. confirmed;<br><br>5) Nursing home resident with symptoms of respiratory infection AND suspicion of a respiratory infection epidemic in the facility | Italy, Tyrol<br><br>From 16 March onwards, all non-essential travel to and from Finland is prohibited |
| March 22nd - April 13th 2020 | Same as 15 March<br>and 6) health care personnel with symptoms of respiratory infection | No designated epidemic areas; all travel considered a risk |
| April 13th 2020 onwards | 1) All individuals experiencing either respiratory or gastrointestinal symptoms;<br><br>2) Patients belonging to risk groups that are admitted to hospital for any reason | |

*New areas in cursive

**eTable 2.** List of COVID-19 cohort wards. Inpatients were admitted to these COVID-19 cohort wards during the study period. Both laboratory confirmed and suspected COVID-19 patients were admitted to the cohort wards, except for two wards (COV_KNKINF and COV_KITEHO) to which only laboratory confirmed cases were admitted.

| Name of the ward | City | Hospital | Type of ward | Date when started as a cohort ward |
|---|---|---|---|---|
| MEKINFOS | Helsinki | Meilahti hospital area | ward | 4.3.2020 |
| MEKINFOSK4B | Helsinki | Meilahti hospital area | ward | 4.3.2020 |
| MEKKEU6A | Helsinki | Meilahti hospital area | ward | 16.3.2020 |
| MEKKEU6B | Helsinki | Meilahti hospital area | ward | 23.3.2020 |
| COV_MEKOS5 | Helsinki | Meilahti hospital area | ward | 1.4.2020 |
| MAOS5 | Helsinki | Malmi Hospital | ward | 13.3.2020 |
| PES4K | Vantaa | Peijas Hospital | ward | 30.3.2020 |
| JOKEU5 | Espoo | Jorvi Hospital | ward | 15.3.2020 |
| HYINFB-4 | Hyvinkää | Hyvinkää Hospital | ward | 8.4.2020 |
| RAUPPAVD | Raasepori | Raasepori Hospital | ward | 23.3.2020 |
| COV_KNKINF | Helsinki | Surgical Hospital | ward | 15.4.2020 |
| COV_KITEHO | Helsinki | Surgical Hospital | ICU | 9.4.2020 |
| MEM1 | Helsinki | Meilahti hospital area | ICU* | ** |
| PET | Vantaa | Peijas Hospital | ICU* | ** |
| PEPV | Vantaa | Peijas Hospital | ICU* | ** |
| JOU2 | Espoo | Jorvi Hospital | ICU* | ** |
| HYTEHVA | Hyvinkää | Hyvinkää Hospital | ICU* | ** |
| POPPKL | Porvoo | Porvoo Hospital | emergency department* | ** |

*Due to lack of patient lists of ICUs or one cohort ward at Porvoo Hospital, all inpatients with SARS-CoV-2 RT-PCR taken at ICUs or at the emergency department of Porvoo Hospital were included.

** All tests taken from 4 March to 15 April included.

**eTable 3**. Estimated SARS-CoV-2 RT-PCR sensitivity values in the laboratory confirmed and high suspicion group combined according to specimen type in the first SARS-CoV-2 RT-PCR test.

| **Specimen type*** | **Inpatients Sensitivity (95 % CI)** | **Outpatients Sensitivity (95 % CI)** | **All Sensitivity (95 % CI)** |
|---|---|---|---|
| Nasopharyngeal | 195/274 | 154/458 | 349/732 |
| | **71.2 %** | **33.6 %** | **47.7 %** |
| | (65.5 – 76.2 %) | (29.4 – 38.1 %) | (44.1 – 51.3 %) |
| Oropharyngeal | 36/54 | 39/97 | 75/151 |
| | **66.7 %** | **40.2 %** | **49.3 %** |
| | (53.4 – 77.8 %) | (31.0 – 50.2 %) | (41.5 – 57.2 %) |
| Not known | 47/84 | 41/117 | 88/201 |
| | **56.0 %** | **35.0 %** | **43.8 %** |
| | (45.3 – 66.1 %) | (27.0 – 44.0 %) | (37.1 – 50.7 %) |

* Other sample types (Brocho-alveolar lavage, sputum, and tracheal aspirate) were excluded from analysis (n=4 for inpatients, n=2 for outpatients). Specimen type was unknown for 84 inpatients and 117 outpatients.

**eTable 4.** Estimated SARS-CoV-2 RT-PCR sensitivity values in the laboratory confirmed, and in the laboratory confirmed and high suspicion group combined according to delay (days) from symptom onset to first SARS-CoV-2 RT-PCR test.

| Delay (days) from symptom onset to sampling | **No of patients with first RT-PCR positive** | **No of COVID-19 Laboratory confirmed patients** | **No of COVID-19 High suspicion patients** | **Total no of COVID-19 Laboratory confirmed and high suspicion patients** | **Sensitivity for COVID-19 Laboratory confirmed patients % (95 % CI)** | **Sensitivity for COVID-19 Laboratory confirmed + High suspicion patients % (95 % CI)** |
|---|---|---|---|---|---|---|
| <1 day | 29 | 37 | 21 | 58 | 78.4 (62.8 - 88.6) | 50.0 (37.5 - 62.5) |
| 1-2 days | 118 | 129 | 132 | 261 | 91.5 (85.4 - 95.2) | 45.2 (39.3 - 51.3) |
| 3-4 days | 96 | 102 | 99 | 201 | 94.1 (87.8 - 97.3) | 47.8 (41.0 - 54.6) |
| 5-6 days | 77 | 88 | 55 | 143 | 87.5 (79.0 - 92.9) | 53.8 (45.7 - 61.8) |
| 7-14 days | 153 | 172 | 136 | 308 | 89.0 (83.4 - 92.8) | 49.7 (44.1 - 55.2) |
| >14 days | 10 | 13 | 26 | 39 | 76.9 (49.7 - 91.8) | 25.6 (14.6 - 41.1) |
| No data | 33 | 33 | 47 | 80 | 100 (89.6 - 100) | 41.2 (31.1 - 52.2) |
| Total | 516 | 574 | 516 | 1090 | 89.9 (87.2 - 92.1) | 47.3 (44.4 - 50.3) |

**eFigure 1.** Number of SARS-CoV-2 RT-PCR tests conducted at the HUSLAB Clinical Microbiology laboratory, and positivity rate over the study period 4 March – 15 April 2020. Median and mean positivity rates were 9.6% and 10%, respectively.

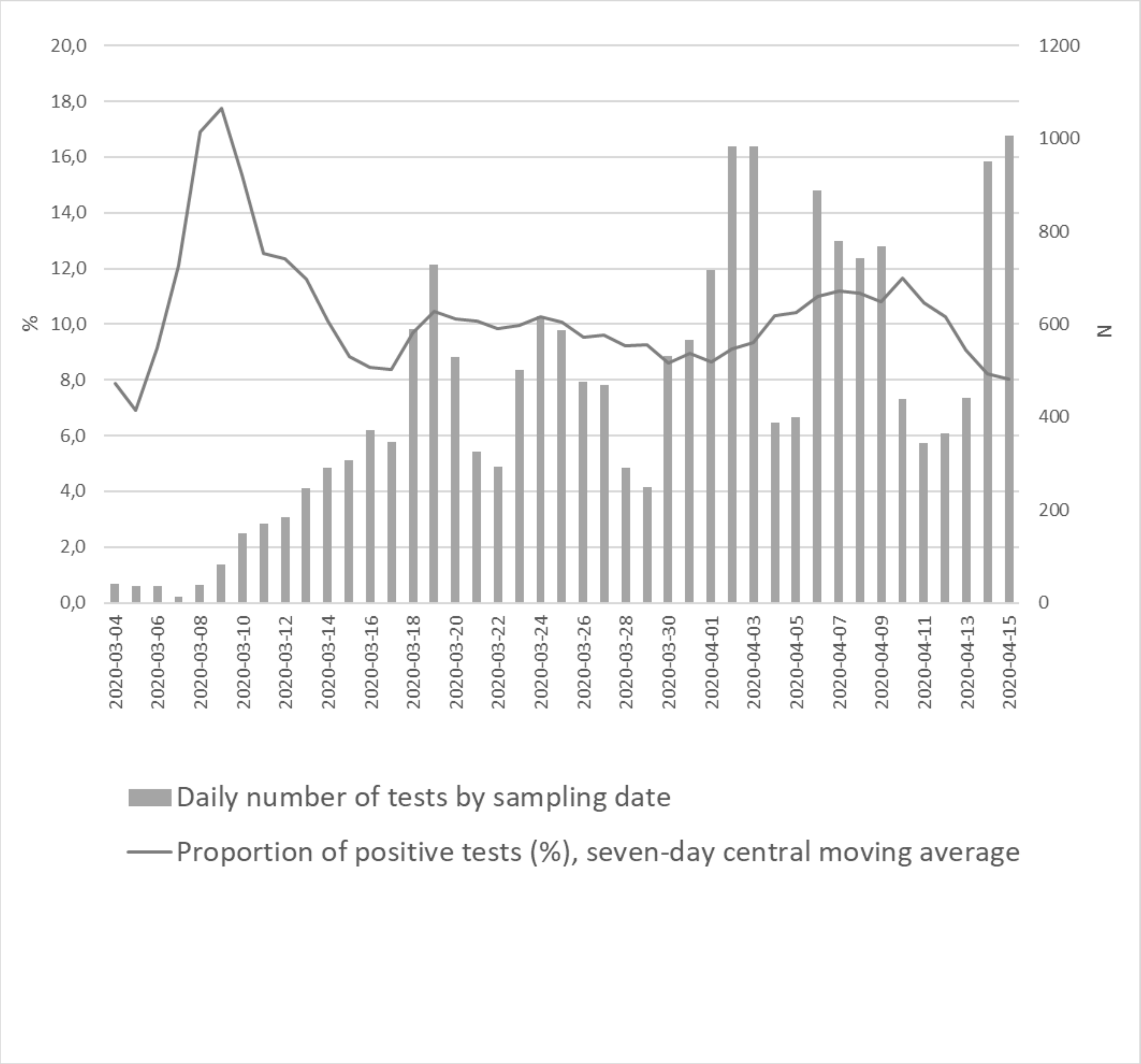

**eFigure 2**. Age distribution of the study population as histograms. A. Inpatients. B. Outpatients.

A

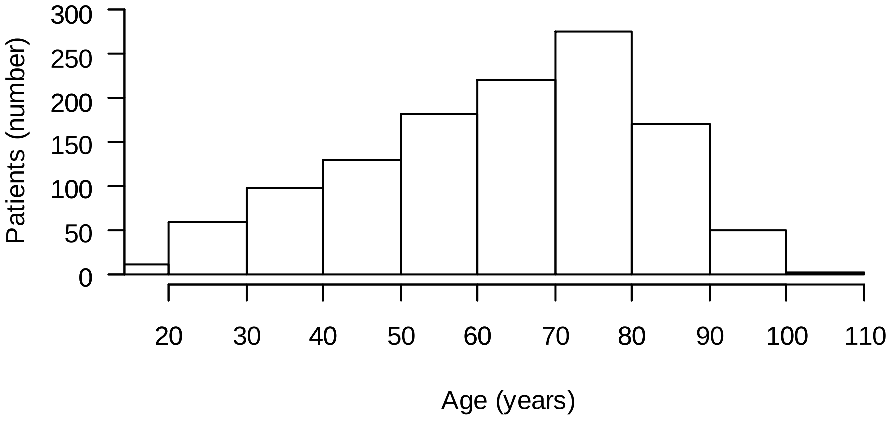


B

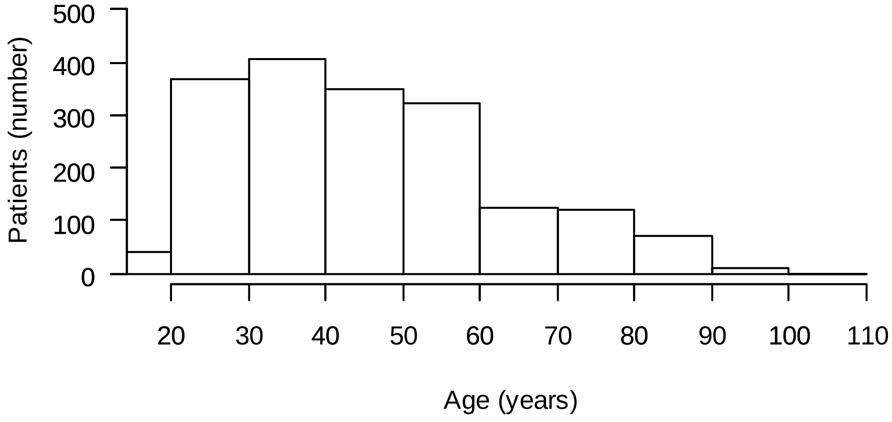

**eFigure 3** Delay to first SARS-CoV2 RT-PCR test (days) in the laboratory confirmed cases and the high suspicion group. A. Inpatients B. Outpatients.

A

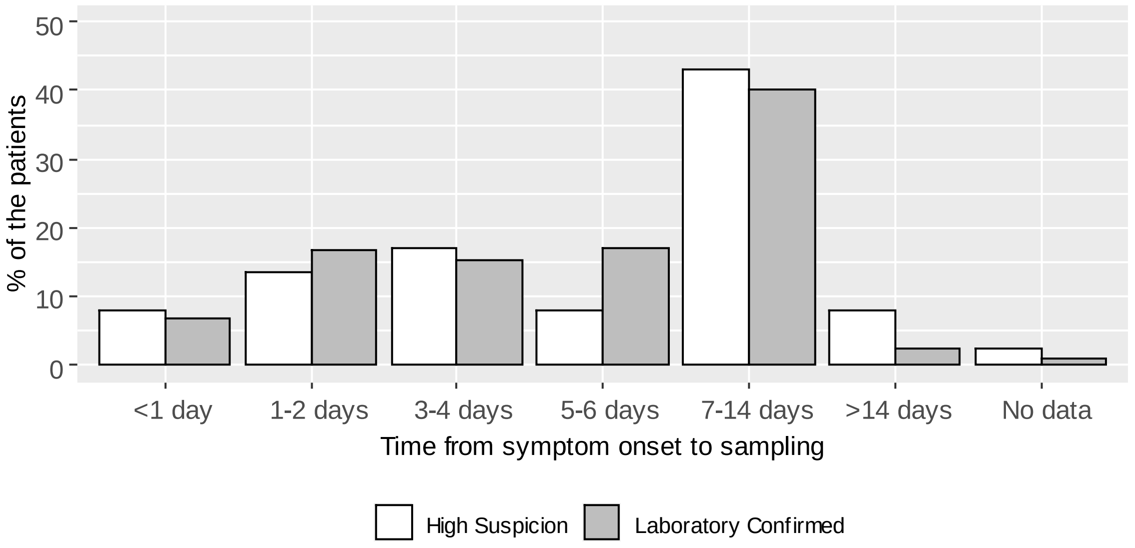


B

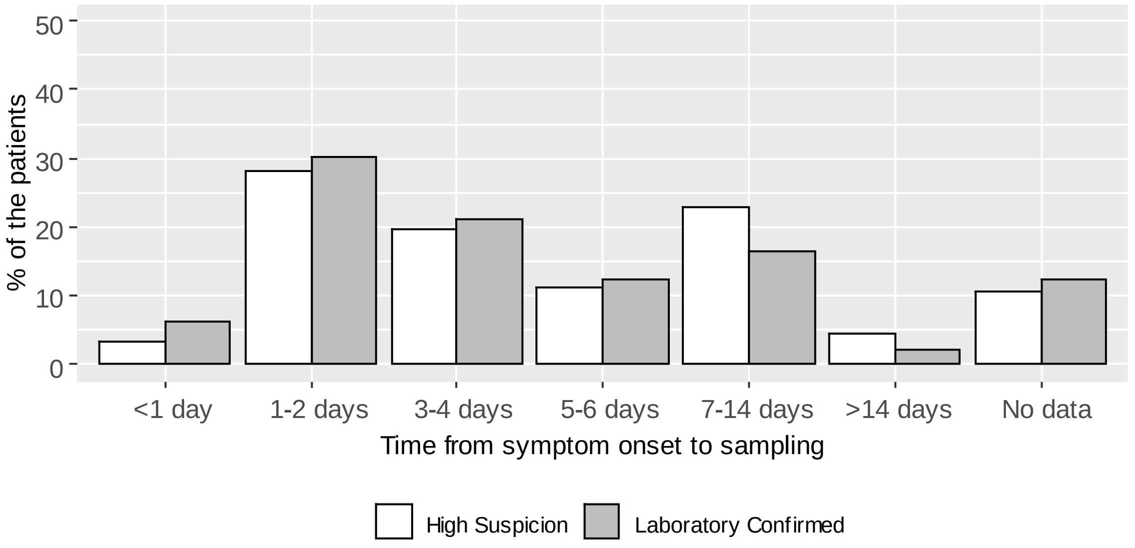

**eFigure 4** Visualisation of clinical sensitivity estimates for inpatients and outpatients according to specimen type.

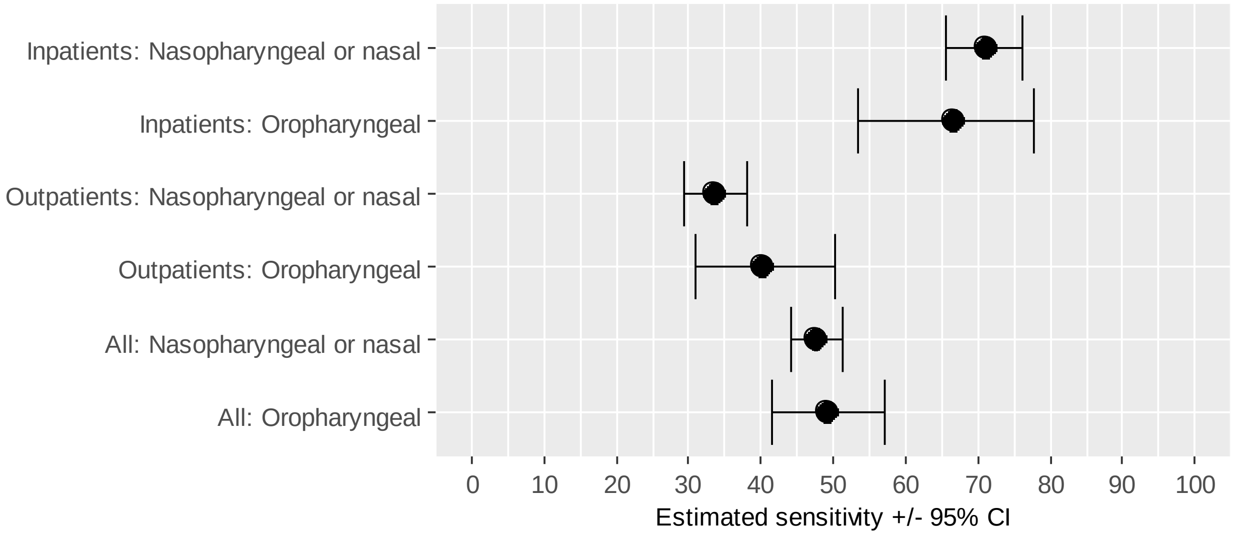
